# Subcortical alterations in newly diagnosed epilepsy and associated changes in brain connectivity and cognition

**DOI:** 10.1101/2024.05.14.24307274

**Authors:** Christophe E de Bézenac, Nicola Leek, Guleed Adan, Rajiv Mohanraj, Shubhabrata Biswas, Anthony G Marson, Simon S Keller

## Abstract

**Objectives:** Patients with chronic focal epilepsy commonly exhibit subcortical atrophy, particularly of the thalamus. The timing of these alterations remains uncertain, though preliminary evidence suggests that observable changes may already be present at diagnosis. It is also not yet known how these morphological changes are linked to the coherence of white matter pathways throughout the brain, or to neuropsychological function often compromised before anti-seizure medication treatment. This study investigates localised atrophy in subcortical regions using surface shape analysis in individuals with newly diagnosed focal epilepsy (NDfE) and assesses their implications on brain connectivity and cognitive function.

**Methods:** We collected structural (T1w) and diffusion weighted MRI, as well as neuropsychological data from 104 patients with NDfE and 45 healthy controls (HC) matched for age, sex and education. A vertex-based shape analysis was performed on subcortical structures to compare patients with NDfE and HC, adjusting for age, sex and intracranial volume. The mean deformation of significance areas (pcor < 0.05) was used to identify white matter pathways associated with overall shape alterations in patients relative to controls using correlational tractography. Additionally, the relationship between significant subcortical shape values and neuropsychological outcomes was evaluated using a generalised canonical correlation approach.

**Results:** Shape analysis revealed bilateral focal inward deformation (a proxy for localised atrophy) in anterior areas of the right and left thalamus and right pallidum in patients with NDfE compared to HC (FWE corrected). No structures showed areas of outward deformation in patients. The connectometry analysis revealed that fractional anisotropy (FA) was positively correlated with thalamic and pallidal shape deformation, i.e., reduced FA was associated with inward deformation in tracts proximal to and or connecting with the thalamus including the fornix, frontal, parahippocampal and corticothalamic pathways. Thalamic and pallidal shape changes were also related to with increased depression and anxiety, and reduced memory and cognitive function.

**Discussion:** These findings suggest that atrophy of the thalamus, which has previously been associated with the generation and maintenance of focal seizures, may present at epilepsy diagnosis and relate to alterations in both white matter connectivity and cognitive performance. We suggest that at least some alterations in brain structure and consequent impact on cognitive and affective processes are the result of early epileptogenic processes rather than exclusively due to the chronicity of longstanding epilepsy, recurrent seizures, and treatment with anti-seizure medication.

## Introduction

Epilepsy is a debilitating neurological condition characterized by unprovoked recurrent seizures that effects around 1% of the population [Fisher et al., 2014]. Seizures result from an imbalance between cortical excitation and inhibition in the brain. Neuroimaging studies into chronic focal epilepsy also commonly identify structural brain alterations using magnetic resonance imaging (MRI). Volume reduction, or atrophy, of temporal lobe structures is a common finding in patients with temporal lobe epilepsy (TLE), typically implicating the hippocampus, entorhinal and perirhinal cortex, and amygdala on the side of the brain where seizures originate [Barron et al., 2013; Bonilha and Keller, 2015]. Subcortical atrophy is also frequently observed in both cerebral hemispheres and is thought to contribute to the generation, maintenance, and spread of epileptic activity [Bonilha and Keller, 2015; Whelan et al., 2018]. The thalamus, in particular, may play a key role in this regard as a complex relay and processing hub of neural signals between the limbic system, the hippocampus, and cortical areas [Kumar et al., 2017].

However, it is not yet clear when epilepsy-related morphometric alterations emerge. Although not all findings are consistent [Salmenperä et al., 2005], preliminary evidence indicates that temporal and subcortical atrophy, particularly in the thalamus, is already present at diagnosis [Kalviainen et al., 1997; Leek et al., 2021]. This suggests that such alterations are related to an early epileptogenic process that may even begin before the manifestation of seizure activity, rather than resulting from prolonged exposure to anti-seizure medication or from the long-term sequelae of repeated seizures [Pohlmann-Eden, 2011]. Given that quantitative neuroimaging methods have highlighted their diagnostic utility in those with established focal epilepsy, it is important to evaluate whether the quantification of subcortical regions can be applied at the earliest reliable clinical timepoint, following a new diagnosis of focal epilepsy for the dual purpose of diagnosis and prognostication.

Studies examining the subcortex, however, tend to use the total volume of each region which can obscure heterogeneous local effects through averaging. In contrast, shape analyses localizes subregional morphometric differences using Bayesian models to estimate surface deformation. While various potential factors such as alterations in myelin, vasculature, and neuronal pruning contribute to localized shape variation [Serbruyns et al., 2015], a growing body of evidence suggests that subregional specificity offers additional insight into pathological brain alterations that conventional volumetric measurements cannot capture [Wang et al., 2022].

There is also widespread agreement that epilepsy is a network disorder [Scharfman et al., 2018], as evidenced by changes in how discrete brain regions structurally and functionally connected to one another [Bernhardt et al., 2015; Richardson, 2012]. Diffusion tensor imaging (DTI) studies in particular have found alterations in the integrity of white matter tracts, which may disrupt the communication between different brain regions and contribute to the development and propagation of seizures [Hatton et al., 2020; Leyden et al., 2015]. Atypical connectivity may already be present in recently-diagnosed patients with epilepsy [Kreilkamp et al., 2021], though detecting these changes is challenging in a population where the localization of the seizure focus is often still unknown. Overall white matter connectivity in the brain has not yet been investigated in relation to morphometric features of brain regions proposed to be susceptible to alterations in focal epilepsy irrespective of where seizures originates (common hubs).

Subcortical structures and, in particular, the thalamus are densely connected to the cortex and plays an important role in motoric, affective, and cognitive functioning [Janacsek et al., 2022]. Neuropsychological processes such as memory and psychomotor speed have been shown to be compromised before the start of antiepileptic drug treatment [Taylor et al., 2010]. However it is not yet clear whether atypical subcortical morphology is associated with cognitive dysfunction in patient with NDfE.

In the present study we examined whether patients with NDfE show evidence of localised subcortical shape alterations compared to neurotypical controls and whether such changes correspond to previously observed patterns of atrophy in the early stages of the disorder. A secondary objective was to examine shape features in relation to whole-brain white matter pathways using correlational tractography, as well as in relation to neuropsychological function using a generalised canonical correlation approach. Through the use of quantitative imaging, the study sought to uncover multivariate patterns of abnormalities at diagnosis with the aim of identifying potential biomarkers of disease progression.

## Methods

### Participants

We investigated 149 participants recruited from 2019 to 2023 as part of the EPINET project: an observational cohort study set up to investigate imaging network markers of cognitive dysfunction and pharmacoresistance in NDfE [Bézenac et al., 2019]. This included 104 patients with a diagnosis of epilepsy of less than 4 months, as well as 45 healthy controls without neurological or psychiatric diagnoses and no family history of epilepsy. Diagnosis of focal epilepsy was determined by expert epileptologists based on history and the assessment of seizure semiology, in accordance with International League Against Epilepsy (ILAE) operational classifications [Fisher et al., 2017]. Patients with known progressive neurological disease, a previous diagnosis of epilepsy, provoked seizures, acute symptomatic seizures, primary generalized seizures or dissociative seizures were not eligible for recruitment. We took the pragmatic decision to not limit recruitment to drug-naïve patients. Additional details relating to the patient cohort can be found in [Bézenac et al., 2024].

### Standard Protocol Approvals, Registrations, and Patient Consents

Written, informed consent was obtained from all participants and the study was approved by the North West, Liverpool East Research Ethics Committee (19/NW/0384) through the Integrated Research Application System (Project ID 260623). Health Research Authority (HRA) approval was provided and the project was sponsored by the UoL (UoL001449) and funded by a UK Medical Research Council (MRC) research grant (MR/S00355X/1).

### MRI acquisition

All patient and control participants were scanned at the Liverpool Magnetic Resonance Imaging Centre (LiMRIC), University of Liverpool, UK. We used a 3T MR system (Siemens Prisma) to acquire a number of MR scans including a 3D T1-weighted (T1w) and Diffusion Kurtosis (DKI) images. A Magnetization Prepared Rapid Gradient Echo (MPRAGE) sequence was used for T1w images (TE = 5.57 ms; TR = 2040 ms; TI = 1,100 ms; slice thickness = 1 mm; voxel size = 1 mm × 1 mm; 176 slices; flip angle = 8).

A multishell diffusion scheme was used for the DKI scan which was acquired in an A>>P phase encoding direction, with a repetition time of 3200 msec and an echo time of 90 msec. In addition to b=0 images (five averages), b-values were 1000 and 2000 s/mm2. The number of diffusion sampling directions were 64 and 64, respectively. The in-plane resolution and slice thickness was 2.5 mm, with a total of 50 slices (no gaps) acquired per participant. To address image distortion, an additional b=0 image with the same dimensions was acquired with opposite phase encoding polarity (P>>A).

### Neurocognitive testing

On the day of scanning, participants also completed a battery of computerized neuropsychological tests shown to highlight cognitive deficits in people with NDfE [Baker et al., 2011; Taylor et al., 2010]. These included assessment tools to evaluate: auditory memory through story recall and recall of verbal pairs (WMS-IV) [Iverson et al., 2013]; visual memory through reproduction of drawings and recall of designs (WMS-IV); working memory and attention through digit span and arithmetic tasks (WAIS-IV) [Baker et al., 2011; Iverson et al., 2013]; processing speed through a coding and symbol search task (WAIS-IV); psychomotor speed through a finger tapping and visual reaction time task; executive functioning through verbal fluency and colour-word interference tasks (D-KEFS) [Scarpina and Tagini, 2017; Swanson, 2005]; mood including depression (PHQ-9) [Fiest et al., 2014] and anxiety (GAD-7) [Seo et al., 2014]; perceived cognitive impairment (ABNAS) [Brooks et al., 2001]; and quality of life (QOLIE-31) [Ridsdale et al., 2017].

### Subcortical shape analysis

The vertex analysis was conducted using FSL’s FIRST (http://fsl.fmrib.ox.ac.uk/fsl/fslwiki/FIRST) to identify variations in the shape of the thalamus. We performed quality control following FSL-FIRST guidelines: no output displayed segmentation errors necessitating manual intervention. Following segmentation, the vertex locations of each participant’s mesh-based representation of the thalamus in both hemispheres were projected onto the surface of the cohort’s average shape transformed to Montreal Neurological Institute space. These projection values indicate the perpendicular distance from the group average surface, with negative values indicating inward deformation and positive values outward deformation in relation to this surface (i.e., growth or atrophy).

Statistical analyses on these scalar projections were carried out using univariate permutation methods through FSL’s randomise tool. Between-group comparisons (NDfE versus Controls) were performed separately for the left and right thalamus using 10000 permutations, controlling for effects of age, sex and intercranial volume. The analysis employed threshold-free cluster enhancement (with 2D opt) corrected for multiple comparisons at an alpha level of 0.05. The mean scalar projection values were extracted for each subject using the fslstats commandline utility for summary statistics and post-hoc testing. Given evidence of group differences found in other subcortical structures, we used the process described above to examine shape differences in other regions segmented though FIRST, including the brainstem and bilateral amygdala, caudate, hippocampus, nucleus accumbens, pallidum, and putamen.

### Connectometry analysis

Diffusion data were first preprocessed with an in-house script which combines MRtrix3 (https://www.mrtrix.org/) and FSL (v.6.0.4) tools for denoising, rician noise filtering and correction for Gibbs ringing and eddy-current induced distortions with TOPUP [Andersson et al., 2003], followed by movement correction with EDDY [Andersson and Sotiropoulos, 2016]. Diffusion MRI connectometry analyses were then carried out using DSI Studio using suggested parameters [Yeh et al., 2016b] (http://dsi-studio.labsolver.org, version Feb 2021). The b-table was checked by an automatic quality control routine to ensure its accuracy [**Sschilling2019?**]. The diffusion data were reconstructed in the MNI space using q-space diffeomorphic reconstruction [Yeh and Tseng, 2011] to obtain the spin distribution function [Yeh et al., 2010]. A diffusion sampling length ratio of 1.25 was used The output resolution of is 2 mm isotorpic. The restricted diffusion was quantified using restricted diffusion imaging [Yeh et al., 2017]. Fractional anisotropy (FA) was extracted as the local connectome fingerprint and used in the connectometry analysis [Yeh et al., 2016b].

Diffusion MRI connectometry was used to derive the correlational tractography that correlates normalized FA with mean deformation values of regions found to be different between patients and controls [Yeh et al., 2016a]. A nonparametric Spearman partial correlation was used to derive correlation values and the effects of age and sex were removed using a multiple regression model. A T-score threshold of 4 was assigned and tracked using a deterministic fiber tracking algorithm to obtain correlational tractography [Yeh et al., 2013]. The tracks were filtered by topology-informed pruning with 16 iterations [Yeh et al., 2019]. A length threshold of 15 voxel distance was used to select tracks. To estimate the false discovery rate, a total of 5000 randomised permutations were applied to the group label to obtain the null distribution of the track length. We also ran a group comparison connectometry analysis (T-score threshold of 2.5, length threshold of 15 voxel distance, 16 iterations, 5000 randomised permutations) to examine tract areas with increased and decreased FA in NDfE compared to HC seeding form regions with significant deformation in patients. A seeding region was placed at left thalamus (45,48,28) right thalamus (33,47,28), right globus pallidus externa (29,39,25), and the right globus pallidus interna (30,41,23).

### Neurocognitive analysis

Relationships between neuropsychological variables and shape deformation in areas displaying group differences was first examined with an exploratory graph analysis using the EGAnet R package [Golino and Epskamp, 2017] to estimates the optimal number of data types/blocks of the full dataset using the graphical lasso and the Louvain algorithm for data-driven community detection. For relationship visualization we then used a Data Integration Analysis for Biomarker discovery using a Latent component method for Omics (DIABLO) implemented in the mixOmics package (v6.20.0) [Rohart et al., 2017], R (v4.2.0) [Team, 2009]. This multivariate integrative method identifies common information across different data types confirmed in the previous exploratory step, while discriminating between groups using a partial least squares discriminant analysis. ROC (Receiver Operating Characteristic) curves and AUC (Area Under the Curve) values are calculated from the dataset for each block as a complementary performance measure based on the predicted scores, with the associated p-value derived from a Wilcoxon test between the predicted scores for one class vs the other.

### Data Availability

The data that support the findings of this study are available via the corresponding author, on reasonable request.

## Results

### Sample characteristics

All 104 patients aged 35.8 ± 13.5 years (range 16–69 years) were included in the analyses, 47 of whom were female (45.2%). In contrast, the 45 controls analysed were aged 40.7 ± 15.7 years (range 17–68 years) including 21 females (56.7%). Age was included as a covariate in all analysis as control were older on average than patients (p=0.052). Controls were also matched for educational attainment: 60% of patients completed high school and 62% of controls (p=0.76) and a comparable percentage identified as white British: 94 patients (90.4%) and 40 controls (88.9%) (p=0.697). Scanning took place an average of 1.3 months (± 1.2, range, 0–3 months) after the commencement of anti-seizure medication treatment – a time interval when we did not expect medication-related changes of brain structure or cognition. Participant demographics, neuroradiological details, and neuropsychology are presented for both patients and controls in a separate publication [Bézenac et al., 2024].

### Subcortical shape results

The shape analysis revealed bilateral focal inward deformation (a proxy for atrophy) in patients with NDfE compared to HC. Significant clusters (TFCE-corrected p<.05) were located in anterior areas of the left thalamus (peak location MNI152 = -15 -6 11; t-statistic = 2.97; pcor = 0.009; number of vertices = 2291) and right thalamus (peak location MNI152 = 4 -7 1; t-statistic = 3.09; pcor = 0.009; number of vertices = 1384). According to the Melbourne Subcortex Atlas (3T group-parcellation) [Tian et al., 2020], peak vertices corresponded to the superior ventroanterior area of the left thalamus (THA-VAs-lh) and the inferior ventroanterior area of the left thalamus (THA-VAia-rh). Apart from the thalamus, the only other subcortical region with significant inward deformation was the right pallidum (peak location MNI152 = 27 -13 -4; regions = THA-VAia-rh; t-statistic = 3.11; pcor = 0.02; number of vertices = 400). The peak location corresponded to the posterior globus pallidus, though significant clusters where found in anterior regions as well. No other regions exhibited cluster-corrected group differences: brain stem (pcor = 0.064), left accumbens (pcor = 0.824), right accumbens (pcor = 0.591), left amygdala (pcor = 0.311), right amygdala (pcor = 0.5), left caudate (pcor = 0.696), right caudate (pcor = 0.61), left hippocampus (pcor = 0.542), right hippocampus (pcor = 0.329), left pallidum (pcor = 0.095), left putamen (pcor = 0.166), and right putamen (pcor = 0.446). No structures showed areas of outward deformation in patients with NDfE compared to controls. Shape related group differences in the right and left thalamus and right pallidum are displayed in Figure 1. Average deformation within these clusters was extracted for each participant and plotted by group: red represents patients, while green represent healthy individuals.

**Fig. 1.**
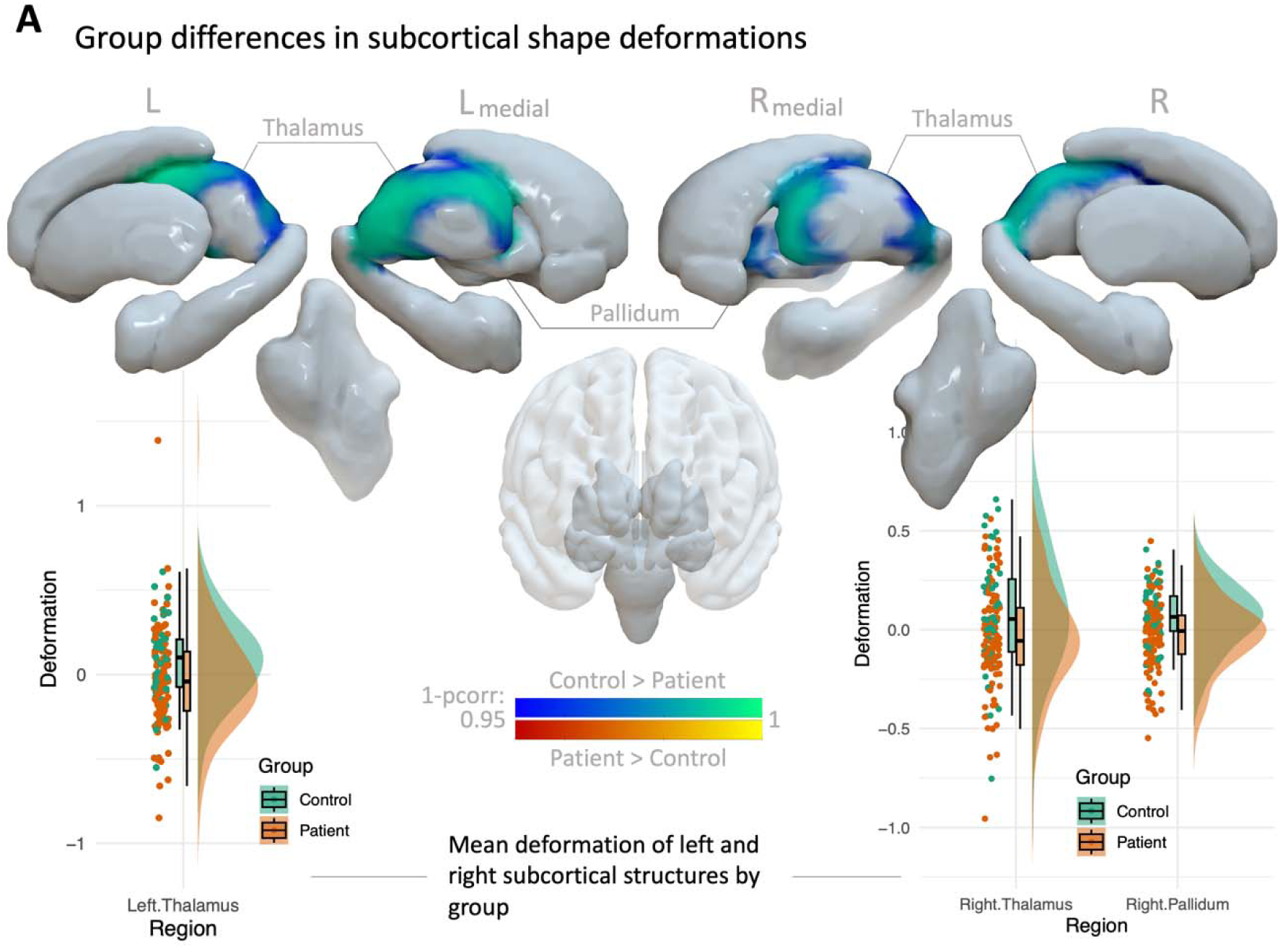
Group differences in subcortical shape deformation. Areas of significant inward shape deformation (blue to turquoise) in the left and right thalamus and right pallidum in patients compared to healthy controls after removing the effects of sex, and intercranial volume. Clusters were identified using threshold-free cluster enhancement (TFCE), corrected to a family-wise error p<.05. No significant outward deformation was found in patients (red to yellow). Average deformation within significant clusters was extracted for each participant and shown by group with box and density plots (red = patients with NDfE; green = control).

### Connectometry results

The whole brain connectometry analysis found tracts with higher FA positively associated with deformation (FDR = 0.000031) in the left and right fornix, corticothalamic pathway, cingulum parolfactory and frontal parahippocampal, dorsal longitudinal fasciculus, as well as inferior-posterior tracts in the cerebellum (Figure 2A). Without defining seed regions, these tract segments centered around superior regions of the thalamus corresponding to where patients with NDfE displayed significant inward deformation compared to controls in the vertex analysis. No tracts showing significant higher FA associated with inward deformation (FDR = 1).

**Fig. 2.**
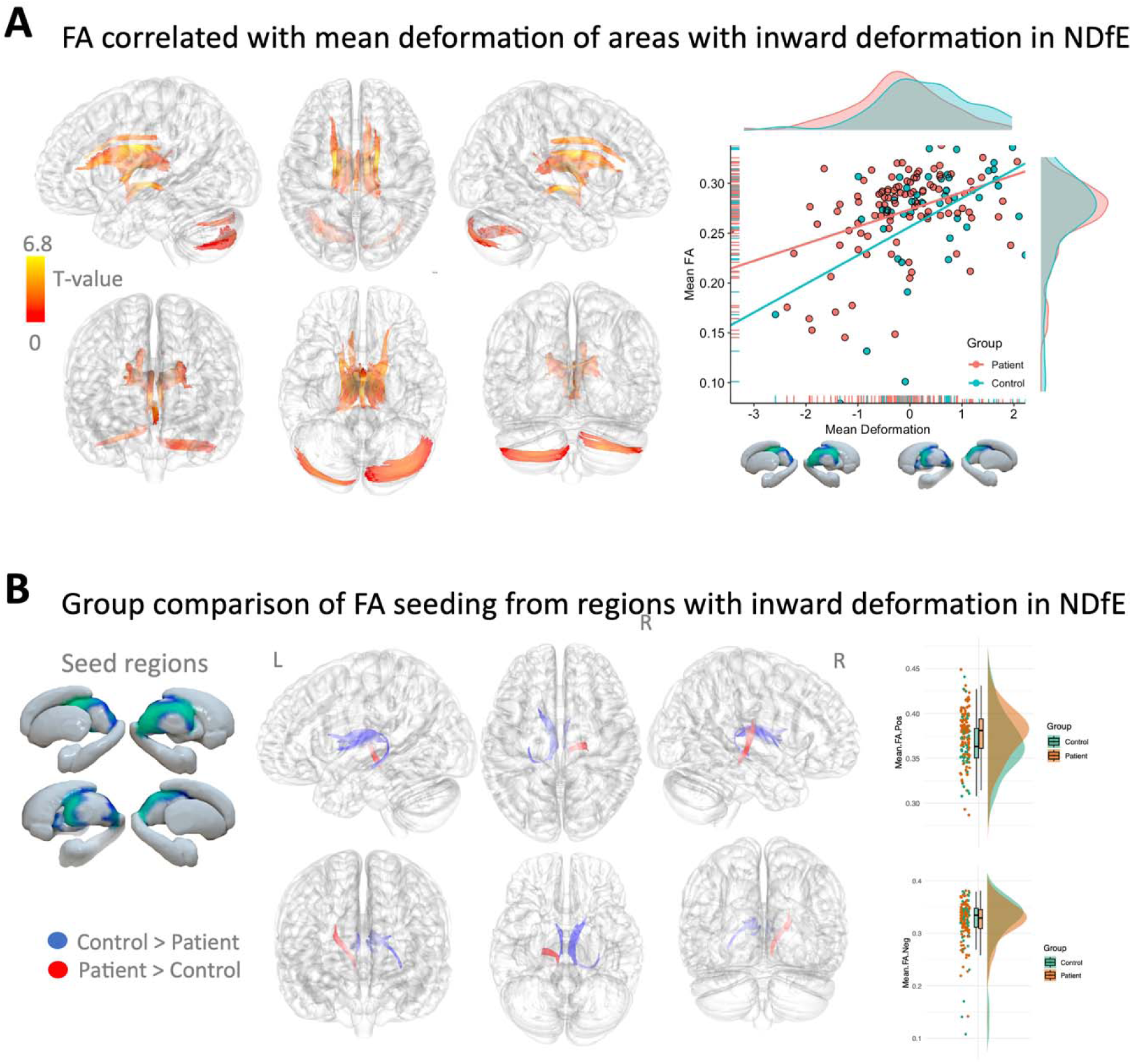
Connectometry results of shape deformation of subcortical areas with inward deformation in patients. (A) White matter tracts with significant positive correlation between FA and shape deformation after controlling for age and sex (red to yellow) (FDR = 0.000031). Scatter and density plots of mean FA across significant tracts for each participant is shown as a function of shape deformation (right). (B) Areas of increased (red) (FDR = 0.000757) and decreased (FDR = 0.017696) FA in patients with NDfE compared to controls (controlling for age and sex) seeding from regions with significant inward and deformation in patients, including the right and left thalamus and right pallidum. Bar and density plots for mean FA in areas of significant positive (top right) and negative (bottom right) FA differences by group. L = left, R = Right, FA = fractional anisotropy, FDR = false discovery rate.

Seeding form regions with significant inward deformation in patients (left and right thalamus and right pallidum), the second connectometry analysis revealed tract segments with increased and decreased FA in patients with NDfE compared to controls (Figure 2B). More specifically, we found reduced FA in patients in parts of the fornix and the part of the corticothalamic tract leading to frontal regions particularly in the left hemisphere (FDR = 0.018). Tract regions with increased FA in patients compared to controls were also found in the right hemisphere in the spinothalamic and corticothalamic tracts involved in sensorimotor processing (FDR = 0.0008).

### Neurocognitive results

The exploratory graph analysis detected five communities which confirmed the internal consistency of the data blocks used in the subsequent DIABLO analysis. The circos plot in Fig. 3 shown the strongest positive (red) and negative (blue) correlations (r>0.8) between variables between the brain data block and all other blocks (edges inside the circle), using components 1 and 2 to integrate the different data types. It also shows the average value of each variable for patients (mustard) and controls (blue) (line profile outside the circle). As the figure indicates, mean deformation in areas of the left and right thalamus was negatively associated with depression scores (PHQ9) and positively correlated with immediate memory. Deformation in the right thalamus additionally shown positive correlation with delayed memory, auditory memory, right hand finger-tapping, and executive function. The right pallidum deformation was negatively related to anxiety and visual reaction time and positively correlated with visual memory and processing speed. We then computed the AUC for each data block as part of the DIABLO analysis. Using components 1 and 2, the mood data block which included anxiety and depression had the highest predictive power with an AUC of 0.84, followed by the cognition (AUC=0.68), brain (AUC=0.66), motor (AUC=0.64), and memory (AUC=0.63) blocks.

**Fig. 3.**
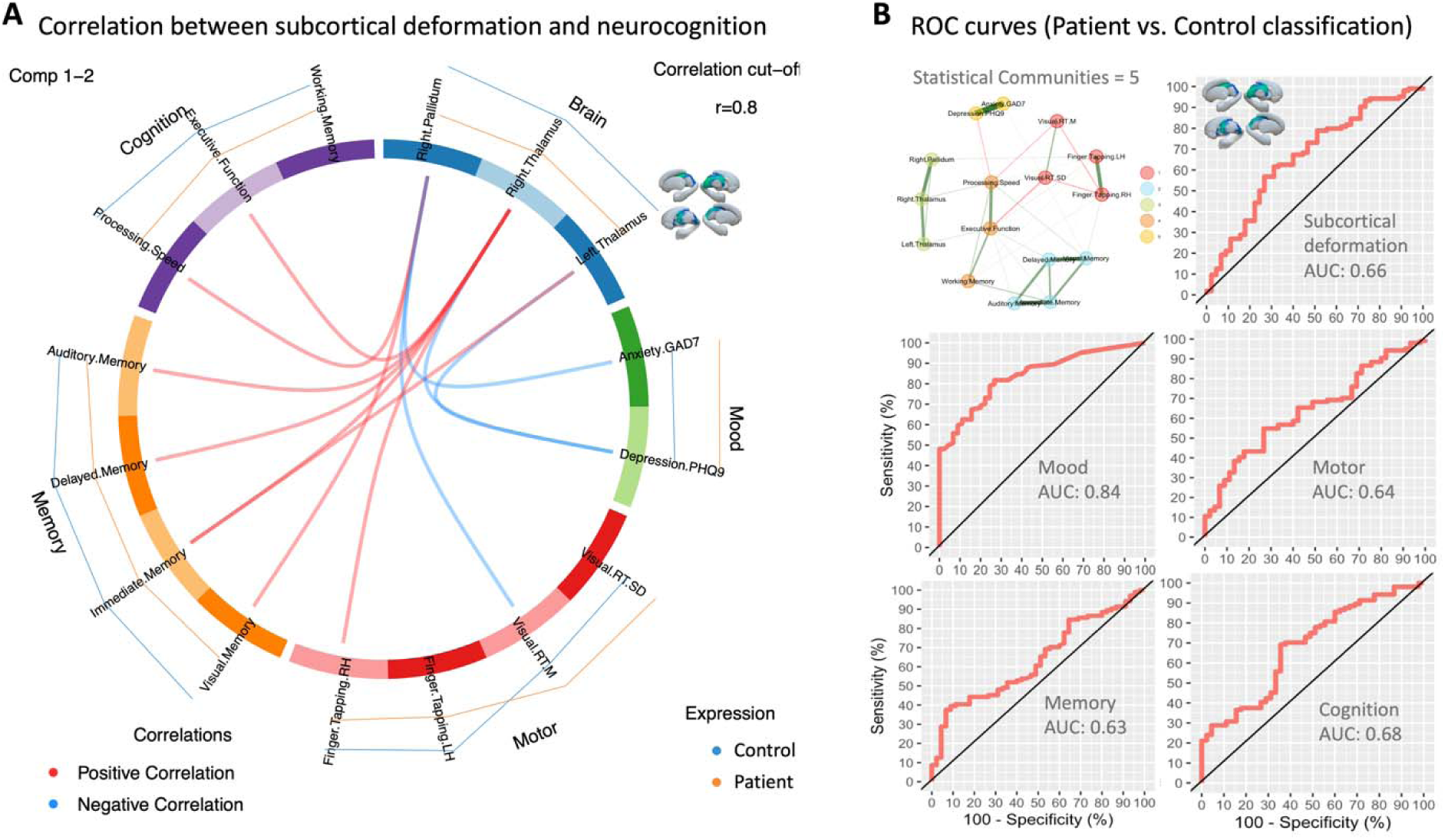
Relationship between subcortical deformation and neuropsychology. (A) Circos plot represents the strongest positive (red) and negative (blue) correlations (> r=0.8) between variables within and between each block (edges inside the circle) and shows the average value of each variable for patients (mustard) and controls (blue) (line profile outside the circle) using components 1 and 2 to integrate the different data types. (B) The exploratory graph network is shown in the top left with node color indicating the 5 detected communities. ROC curves (with AUC values) for distinguishing between patients with NDfE from controls using components 1 and 2 are shown for each data block included in the DIABLO analysis.

## Discussion

The current study examined subcortical structural alterations and their impact on white matter connectivity and neuropsychological performance in a sample of patients with NDfE compared with a group of age, sex and education matched healthy controls. Consistent with our hypothesis based on preliminary work [Kalviainen et al., 1997; Leek et al., 2021], we detected altered surface morphometry of the thalamus bilaterally in patients compared to controls, as well as the right pallidum. No significant group differences of inward or outward deformation were found in other subcortical structures. Furthermore, areas of shape differences between groups correlated with alterations in white matter connectivity as measured by FA derived from diffusion-weighted MRI in a number of tracts proximal to and or connecting with the thalamus, including the fornix and corticothalamic tracts. Subcortical structural changes were also related to measures of anxiety, depression, memory and cognitive function. To our knowledge, this is the first report of interrelated brain structural, connectivity, and cognitive impairments at the point of epilepsy diagnosis.

The association between inward shape deformation in both the left and right thalamus in patients with NDfE supports existing literature, indicating frequent volume loss in both hemispheres of the thalamus in patients with various chronic focal and generalized epilepsies [Anon, 2015; Whelan et al., 2018]. Given its role as a complex relay and processing hub of neural signals between the limbic system, hippocampus, and cortical areas, it makes biological sense that the thalamus is crucial for generating, maintaining, and spreading epileptic activity [Kumar et al., 2017]. It is increasingly seen as an integral part of the neural circuitry of limbic epilepsy with growing evidence that local neuronal changes can increase seizure-related excitability of thalamolimbic circuits [Anon, 2015; Bertram et al., 2001]. For example, a stereoelectroencephalography (SEEG) study found increased synchrony between the thalamus and temporal lobe regions during seizures [Guye et al., 2006], and electrical stumulation of the medial pulvinar nucleus reduced seizure severity in patients with drug-resistent TLE, possibly through the disruption of corticothalamic coupling [Homman-Ludiye and Bourne, 2019]. According to a meta-analysis of voxel-based morphometry studies in refractory TLE, thalamic atrophy is nearly as prevalent as hippocampal atrophy in this cohort [Barron et al., 2013].

Our study is more specifically in line with preliminary findings that show reduced volume of the thalamus from early epilepsy [Leek et al., 2021]. Another study that compared a small samples of patients with newly diagnosed and chronic epilepsy to healthy controls found a trend towards reduced volume in the left and right thalamus in early epilepsy, with left thalamic volume also negatively correlated with disease duration [Park et al., 2015]. Taken together, these findings suggest that observed thalamic changes are likely to develop prior to the manifestation of habitual epilepsy, perhaps before hippocampal volume loss in TLE cases, with additional damage occurring for some as the disorder progresses [Kälviäinen and Salmenperä, 2002].

The shape based approach adopted in our study is more sensitive to local variation than previous investigations into early epilepsy, allowing the identification of regional effects that could be masked by averaging across entire structures. This is particularly relevant to examining structurally and functionally complex areas like the thalamus. In our results, the peak location of inward deformation corresponded to the ventral anterior parts of the thalamus in both hemispheres. Research has shown that high-frequency stimulation of the anterior nucleus of the thalamus can desynchronize the epileptic network in patients with drug-resistant epilepsy and result in a 69% reduction in seizure frequency [Salanova et al., 2015]. It may be that this part of the thalamus plays a particularly important role in regulating seizure-related network dynamics and is increasingly proving to be an effective neuromodulatory treatment target [Krishna et al., 2016; Salanova et al., 2015; Yu et al., 2018].

In contrast to some preliminary work [Leek et al., 2021], we did not find group differences in the left or right hippocampus in patients with recently-diagnosed epilepsy - a region typically associated with drug-resistent TLE [Barron et al., 2013; Bonilha and Keller, 2015]. Our findings are however in accordance with another larger prospective MRI study into recently-diagnosed focal epilepsy [Salmenperä et al., 2005]. It may be that larger samples are also more heterogeneous, including patients with both left and right lateralized, lesional and nonlesional, temporal and frontal lobe epilepsies, which may mask differences only present in a subset of patients. This also leaves open the possibility that changes in the thalamus, in particular, are more closely associated with a mechanism common to different manifestations of focal epilepsy than other subcortical regions and may precede hippocampal changes in the case of chronic TLE.

In addition to thalamic findings, we also found inward deformation in parts of the right pallidum with a trend in the left in patients compared to controls. While there is limited evidence for the direct involvement of this region in seizure generation, there are findings that indicate that the pallidum plays an inhibitory role in modulating seizure activity as part of the basal ganglia network [Deransart et al., 1999], with increased firing rate specifically identified during focal motor seizures [Devergnas et al., 2012]. Quantitative susceptibility mapping has revealed microstructural changes in the globus pallidus in temporal lobe epilepsy [Kiersnowski et al., 2023], while findings from diffusion-tensor imaging data indicate increased centrality of the globus pallidus in patients compared to controls, suggesting its potential role as a network hub, alongside the thalamus, in early focal epilepsy [Park et al., 2019].

Our connectometry results showed that structural alterations of subcortical regions in patients was positively correlated with FA in tracts including the left and right fornix, corticothalamic pathway, cingulum parolfactory and frontal parahippocampal, dorsal longitudinal fasciculus, in addition to inferior-posterior tracts of the cerebellum. The direction of findings suggest that atrophy is associated with reduced FA in tracts proximal to and connecting with the thalamus, despite the fact that we did not introduce seed regions in this analysis. Highlighted tracts are integral components of Papez’ circuit, implicated in both episodic memory consolidation and emotion processing [Kamali et al., 2023], and thought to play a role in seizure propagation [Gologorsky and Alterman, 2011]. While the covariance between brain volume reduction and white matter microstructural integrity makes biological sense [Appelman et al., 2009], it remains uncertain in relation to focal epilepsy whether it is primarily driven by gray matter neurodegeneration or a consequence of changes to white matter architecture.

It is also noteworthy that areas of significant correlation connect to temporal and frontal parts of the brain that are particularly susceptible to developing epileptogenic lesions [Chowdhury et al., 2021] and essential for normal neurocognitive function [Stretton and Thompson, 2012]. Interestingly, the group comparison seeding from regions with significant inward deformation in patients also found reduced FA in the fornix and parts of the corticothalamic tract connected to the frontal lobe, particularly in the left hemisphere. This is in accordance with a recent study that found white matter alterations in thalamic and limbic connections including the fornix, using fixel-based approach in MR-negative patients with temporal and frontal lobe epilepsy [Bartoňová et al., 2023]. The finding of increased FA in patients compared to controls in right spinothalamic and corticothalamic tracts is more difficult to contextualize, though the involvement of these tracts with pain and sensorimotor processing may points towards a compensatory effect of epilepsy-related alterations found in seed regions [Willis and Westlund, 1997].

One of the strengths of our data is the extensive neuropsychological measures which can be examined in relation to neuroimaging features. Here we found that shape deformation in the right and left thalamus was negatively associated with depression and positively related to immediate memory. Recent investigations have proposed a connection between the anterior thalamus and emotional processing in treatment-resistant epilepsy, with bilateral deep brain stimulation targeting this region inducing reversible depression symptoms in patients [Tröster et al., 2017]. Another study showed that damage to the anterior thalamus was also associated with increased depression and anxiety [Scharf et al., 2023]. This points towards a potential role of the anterior thalamus in emotional regulation for patients with epilepsy. Future research could examine common underlying mechanisms between mood disorders and focal epilepsy [Salpekar, 2016], supported by our finding that anxiety and depression were the most effective features for distinguishing patients from controls (AUC=0.84).

We found that structural alterations particularly in the right thalamus were positively associated with memory (delayed and auditory) and executive function. Anterior thalamic nuclei form a key anatomical hub within hippocampal-diencephalic-cortical network that supports memory function [Barnett et al., 2021] and has previously been implicated in focal epilepsy [Whelan et al., 2018]. Reduced thalamic volume, specifically in the anterior and medial nuclei, has been linked to cognitive decline in aging and diminished executive functions, encompassing speed of information processing, focused attention, and working memory [Fama and Sullivan, 2015]. These are domains known to be compromised at diagnosis in patients with focal epilepsy [Taylor et al., 2010]. Our findings also showed that deformation in the pallidum was negatively related to anxiety and visual reaction time and positively correlated with visual memory and processing speed, in accordance with research identifying the role the region plays in the reward circuitry [Smith et al., 2009].

One of the main challenges of studying newly-diagnosed epilepsy is patient heterogeneity. Given that the epileptogenic focus in nonlesional cases is not usually known at diagnosis, the sample is likely to consist of a combination of right and left lateralized, temporal and frontal lobe epilepsy cases. Typically, further investigations that allow sub-grouping are reserved for patients that continue to experience seizures after prolonged anti-seizure medication (ASM) treatment, aiming for more precise localization of seizure focus through semiological analysis, detailed imaging, EEG, and neuropsychological evaluation. We propose, however, that what is lost by not having a highly phenotyped group is gained through a pragmatic approach to studying all patients with a new diagnosis of focal epilepsy. Identifying commonalities, or network hubs such as the thalamus, that apply more generally to focal epilepsy can provide valuable insights into disease mechanisms. However the strengths of both approaches should be combined through longitudinal imaging and neurocognition, with the aim of determining the extent to which clinical features and treatment outcome can be predicted from diagnosis to allow early patient stratification.

In conclusion, our findings highlight the presence of early subcortical brain changes, particularly involving the thalamus, in focal epilepsy, which may not solely be attributed to the chronicity of the condition. The observed thalamic atrophy appears to be associated with alterations in broader structural networks, potentially elucidating the widespread neuropsychological deficits observed in the early stages of epilepsy. Moving forward, it will be important for future research to prioritize the acquisition of longitudinal imaging and cognitive data from the point of diagnosis. This will allow for a more comprehensive mapping of patient-specific clinical, biological, and cognitive trajectories, ultimately enhancing our understanding of epilepsy progression and informing tailored therapeutic interventions to improve outcomes for individuals living with epilepsy.

## Data Availability

All data produced in the present study are available upon reasonable request to the authors

## Notes

### Competing Interest Statement

The authors have declared no competing interest.

### Author Declarations

the study was approved by the North West, Liverpool East Research Ethics Committee (19/NW/0384) through the Integrated Research Application System (Project ID 260623). Health Research Authority (HRA) approval was provided and the project was sponsored by the UoL (UoL001449).

## References

Andersson JL, Skare S, Ashburner J (2003): How to correct susceptibility distortions in spin-echo echo-planar images: Application to diffusion tensor imaging. Neuroimage 20:870–888.

Andersson JL, Sotiropoulos SN (2016): An integrated approach to correction for off-resonance effects and subject movement in diffusion MR imaging. Neuroimage 125:1063–1078.

Thalamotemporal alteration and postoperative seizures in temporal lobe epilepsy (2015): Annals of Neurology 77:760–774.

Appelman AP, Exalto LG, Van Der Graaf Y, Biessels GJ, Mali WP, Geerlings MI (2009): White matter lesions and brain atrophy: More than shared risk factors? A systematic review. Cerebrovascular Diseases 28:227–242.

Baker GA, Taylor J, Aldenkamp AP, group S (2011): Newly diagnosed epilepsy: Cognitive outcome after 12 months. Epilepsia 52:1084–1091.

Barnett S, Parr-Brownlie L, Perry B, Young C, Wicky H, Hughes S, McNaughton N, Dalrymple-Alford J (2021): Anterior thalamic nuclei neurons sustain memory. Current Research in Neurobiology 2:100022.

Barron DS, Fox PM, Laird AR, Robinson JL, Fox PT (2013): Thalamic medial dorsal nucleus atrophy in medial temporal lobe epilepsy: A VBM meta-analysis. NeuroImage: Clinical 2:25–32.

Bartoňová M, Tournier J-D, Bartoň M, Říha P, Vojtíšek L, Mareček R, Doležalová I, Rektor I (2023): White matter alterations in MR-negative temporal and frontal lobe epilepsy using fixel-based analysis. Scientific Reports 13:19.

Bernhardt BC, Hong S-J, Bernasconi A, Bernasconi N (2015): Magnetic resonance imaging pattern learning in temporal lobe epilepsy: Classification and prognostics. Annals of neurology 77:436–446.

Bertram EH, Mangan PS, Zhang D, Scott CA, Williamson JM (2001): The midline thalamus: Alterations and a potential role in limbic epilepsy. Epilepsia 42:967–978.

Bézenac C de, Leek N, Adan G, Ali A, Mohanraj R, Biswas S, Mcginty R, Murphy K, Malone H, Baker G, Moore P, Marson AG, Keller SS (2024): Prospective neuroimaging and neuropsychological evaluation in adults with newly diagnosed focal epilepsy. medRxiv. https://www.medrxiv.org/content/early/2024/05/14/2024.05.14.24307267.

Bézenac C de, Garcia-Finana M, Baker G, Moore P, Leek N, Mohanraj R, Bonilha L, Richardson M, Marson AG, Keller S (2019): Investigating imaging network markers of cognitive dysfunction and pharmacoresistance in newly diagnosed epilepsy: A protocol for an observational cohort study in the UK. BMJ open 9:e034347.

Bonilha L, Keller SS (2015): Quantitative MRI in refractory temporal lobe epilepsy: Relationship with surgical outcomes. Quantitative Imaging in Medicine and Surgery 5:204.

Brooks J, Baker GA, Aldenkamp AP (2001): The a–b neuropsychological assessment schedule (ABNAS): The further refinement of a patient-based scale of patient-perceived cognitive functioning. Epilepsy research 43:227–237.

Chowdhury FA, Silva R, Whatley B, Walker MC (2021): Localisation in focal epilepsy: A practical guide. Practical Neurology 21:481–491.

Deransart C, Riban V, Lê B-T, Hechler V, Marescaux C, Depaulis A (1999): Evidence for the involvement of the pallidum in the modulation of seizures in a genetic model of absence epilepsy in the rat. Neuroscience letters 265:131–134.

Devergnas A, Piallat B, Prabhu S, Torres N, Louis Benabid A, David O, Chabardès S (2012): The subcortical hidden side of focal motor seizures: Evidence from micro-recordings and local field potentials. Brain 135:2263–2276.

Fama R, Sullivan EV (2015): Thalamic structures and associated cognitive functions: Relations with age and aging. Neuroscience & Biobehavioral Reviews 54:29–37.

Fiest KM, Patten SB, Wiebe S, Bulloch AG, Maxwell CJ, Jetté N (2014): Validating screening tools for depression in epilepsy. Epilepsia 55:1642–1650.

Fisher RS, Cross JH, French JA, Higurashi N, Hirsch E, Jansen FE, Zuberi SM (2017): Operational classification of seizure types by the international league against epilepsy: Position paper of the ILAE commission for classification and terminology. Epilepsia 58:522–530.

Fisher RS, Acevedo C, Arzimanoglou A, Bogacz A, Cross JH, Elger CE, Engel Jr J, Forsgren L, French JA, Glynn M, others (2014): ILAE official report: A practical clinical definition of epilepsy. Epilepsia 55:475–482.

Golino HF, Epskamp S (2017): Exploratory graph analysis: A new approach for estimating the number of dimensions in psychological research. PloS one 12:e0174035.

Gologorsky Y, Alterman R (2011): Cerebral–deep. Essential Neuromodulation:47.

Guye M, Régis J, Tamura M, Wendling F, Gonigal AM, Chauvel P, Bartolomei F (2006): The role of corticothalamic coupling in human temporal lobe epilepsy. Brain 129:1917–1928.

Hatton SN, Huynh KH, Bonilha L, Abela E, Alhusaini S, Altmann A, Alvim MK, Balachandra AR, Bartolini E, Bender B, others (2020): White matter abnormalities across different epilepsy syndromes in adults: An ENIGMA-epilepsy study. Brain 143:2454–2473.

Homman-Ludiye J, Bourne JA (2019): The medial pulvinar: Function, origin and association with neurodevelopmental disorders. Journal of anatomy 235:507–520.

Iverson GL, Holdnack JA, Lange RT (2013): Using the WAIS–IV/WMS–IV/ACS following moderate-severe traumatic brain injury. In: WAIS-IV, WMS-IV, and ACS. Elsevier. pp 485–544.

Janacsek K, Evans TM, Kiss M, Shah L, Blumenfeld H, Ullman MT (2022): Subcortical cognition: The fruit below the rind. Annual Review of Neuroscience 45:361–386.

Kalviainen R, Partanen K, Aikia M, Mervaala E, Vainio P, Riekkinen P, Pitkanen A (1997): MRI-based hippocampal volumetry and t sub 2 relaxometry: Correlation to verbal memory performance in newly diagnosed epilepsy patients with left-sided temporal lobe focus. Neurology 48:286–287.

Kälviäinen R, Salmenperä T (2002): Do recurrent seizures cause neuronal damage? A series of studies with MRI volumetry in adults with partial epilepsy. Progress in brain research 135:279–295.

Kamali A, Milosavljevic S, Gandhi A, Lano KR, Shobeiri P, Sherbaf FG, Sair HI, Riascos RF, Hasan KM (2023): The cortico-limbo-thalamo-cortical circuits: An update to the original papez circuit of the human limbic system. Brain topography 36:371–389.

Kiersnowski OC, Winston GP, Caciagli L, Biondetti E, Elbadri M, Buck S, Duncan JS, Thornton JS, Shmueli K, Vos SB (2023): Quantitative susceptibility mapping identifies hippocampal and other subcortical grey matter tissue composition changes in temporal lobe epilepsy. Human Brain Mapping 44:5047–5064.

Kreilkamp BA, McKavanagh A, Alonazi B, Bryant L, Das K, Wieshmann UC, Marson AG, Taylor PN, Keller SS (2021): Altered structural connectome in non-lesional newly diagnosed focal epilepsy: Relation to pharmacoresistance. NeuroImage: Clinical 29:102564.

Krishna V, King NKK, Sammartino F, Strauss I, Andrade DM, Wennberg RA, Lozano AM (2016): Anterior nucleus deep brain stimulation for refractory epilepsy: Insights into patterns of seizure control and efficacious target. Neurosurgery 78:802–811.

Kumar VJ, Oort E van, Scheffler K, Beckmann CF, Grodd W (2017): Functional anatomy of the human thalamus at rest. Neuroimage 147:678–691.

Leek NJ, Neason M, Kreilkamp B, Bézenac C de, Ziso B, Elkommos S, Das K, Marson AG, Keller SS (2021): Thalamohippocampal atrophy in focal epilepsy of unknown cause at the time of diagnosis. European journal of neurology 28:367–376.

Leyden KM, Kucukboyaci NE, Puckett OK, Lee D, Loi RQ, Paul B, McDonald CR (2015): What does diffusion tensor imaging (DTI) tell us about cognitive networks in temporal lobe epilepsy? Quantitative imaging in medicine and surgery 5:247.

Park KM, Han YH, Kim TH, Mun CW, Shin KJ, Ha SY, Park J, Hur YJ, Kim HY, Park SH, others (2015): Cerebellar white matter changes in patients with newly diagnosed partial epilepsy of unknown etiology. Clinical neurology and neurosurgery 138:25–30.

Park KM, Lee BI, Shin KJ, Ha SY, Park J, Kim SE, Kim SE (2019): Pivotal role of subcortical structures as a network hub in focal epilepsy: Evidence from graph theoretical analysis based on diffusion-tensor imaging. Journal of Clinical Neurology (Seoul, Korea) 15:68.

Pohlmann-Eden B (2011): Conceptual relevance of new-onset epilepsy. Epilepsia 52:1–6.

Richardson MP (2012): Large scale brain models of epilepsy: Dynamics meets connectomics. Journal of Neurology, Neurosurgery & Psychiatry 83:1238–1248.

Ridsdale L, Wojewodka G, Robinson E, Landau S, Noble A, Taylor S, Richardson M, Baker G, Goldstein LH (2017): Characteristics associated with quality of life among people with drug-resistant epilepsy. Journal of neurology 264:1174–1184.

Rohart F, Gautier B, Singh A, Lê Cao K-A (2017): mixOmics: An r package for ‘omics feature selection and multiple data integration. PLoS computational biology 13:e1005752.

Salanova V, Witt T, Worth R, Henry TR, Gross RE, Nazzaro JM, Labar D, Sperling MR, Sharan A, Sandok E, others (2015): Long-term efficacy and safety of thalamic stimulation for drug-resistant partial epilepsy. Neurology 84:1017–1025.

Salmenperä T, Könönen M, Roberts N, Vanninen R, Pitkänen A, Kälviäinen R (2005): Hippocampal damage in newly diagnosed focal epilepsy: A prospective MRI study. Neurology 64:62–68.

Salpekar J (2016): Mood disorders in epilepsy. Focus 14:465–472.

Scarpina F, Tagini S (2017): The stroop color and word test. Frontiers in psychology 8:557.

Scharf A-C, Gronewold J, Eilers A, Todica O, Hermann DM (2023): Depression and anxiety in acute ischemic stroke involving the anterior but not paramedian or inferolateral thalamus. Frontiers in psychology 14:1218526.

Scharfman HE, Kanner AM, Friedman A, Blümcke I, Crocker CE, Cendes F, Diaz-Arrastia R, Förstl H, Fenton AA, Grace AA, others (2018): Epilepsy as a network disorder (2): What can we learn from other network disorders such as dementia and schizophrenia, and what are the implications for translational research? Epilepsy & Behavior 78:302–312.

Seo J-G, Cho YW, Lee S-J, Lee J-J, Kim J-E, Moon H-J, Park S-P (2014): Validation of the generalized anxiety disorder-7 in people with epilepsy: A MEPSY study. Epilepsy & Behavior 35:59–63.

Serbruyns L, Leunissen I, Huysmans T, Cuypers K, Meesen RL, Ruitenbeek P van, Sijbers J, Swinnen SP (2015): Subcortical volumetric changes across the adult lifespan: Subregional thalamic atrophy accounts for age-related sensorimotor performance declines. Cortex 65:128–138.

Smith KS, Tindell AJ, Aldridge JW, Berridge KC (2009): Ventral pallidum roles in reward and motivation. Behavioural brain research 196:155–167.

Stretton J, Thompson P (2012): Frontal lobe function in temporal lobe epilepsy. Epilepsy research 98:1–13.

Swanson J (2005): The delis-kaplan executive function system: A review. Canadian Journal of School Psychology 20:117–128.

Taylor J, Kolamunnage-Dona R, Marson AG, Smith PE, Aldenkamp AP, Baker GA (2010): Patients with epilepsy: Cognitively compromised before the start of antiepileptic drug treatment? Epilepsia 51:48–56.

Team RDC (2009): A language and environment for statistical computing. http://www.R-project.org.

Tian Y, Margulies DS, Breakspear M, Zalesky A (2020): Topographic organization of the human subcortex unveiled with functional connectivity gradients. Nature neuroscience 23:1421–1432.

Tröster AI, Meador KJ, Irwin CP, Fisher RS, Group SS, others (2017): Memory and mood outcomes after anterior thalamic stimulation for refractory partial epilepsy. Seizure 45:133–141.

Wang Z, Fontaine M, Cyr M, Rynn MA, Simpson HB, Marsh R, Pagliaccio D (2022): Subcortical shape in pediatric and adult obsessive-compulsive disorder. Depression and anxiety 39:504–514.

Whelan CD, Altmann A, Botía JA, Jahanshad N, Hibar DP, Absil J, Alhusaini S, Alvim MK, Auvinen P, Bartolini E, others (2018): Structural brain abnormalities in the common epilepsies assessed in a worldwide ENIGMA study. Brain 141:391–408.

Willis WD, Westlund K (1997): Neuroanatomy of the pain system and of the pathways that modulate pain. Journal of Clinical Neurophysiology 14:2–31.

Yeh F-C, Badre D, Verstynen T (2016a): Connectometry: A statistical approach harnessing the analytical potential of the local connectome. Neuroimage 125:162–171.

Yeh F-C, Liu L, Hitchens TK, Wu YL (2017): Mapping immune cell infiltration using restricted diffusion MRI. Magnetic resonance in medicine 77:603–612.

Yeh F-C, Panesar S, Barrios J, Fernandes D, Abhinav K, Meola A, Fernandez-Miranda JC (2019): Automatic removal of false connections in diffusion MRI tractography using topology-informed pruning (TIP). Neurotherapeutics 16:52–58.

Yeh F-C, Tseng W-YI (2011): NTU-90: A high angular resolution brain atlas constructed by q-space diffeomorphic reconstruction. Neuroimage 58:91–99.

Yeh F-C, Verstynen TD, Wang Y, Fernández-Miranda JC, Tseng W-YI (2013): Deterministic diffusion fiber tracking improved by quantitative anisotropy. PloS one 8:e80713.

Yeh F-C, Vettel JM, Singh A, Poczos B, Grafton ST, Erickson KI, Tseng W-YI, Verstynen TD (2016b): Quantifying differences and similarities in whole-brain white matter architecture using local connectome fingerprints. PLoS computational biology 12:e1005203.

Yeh F-C, Wedeen VJ, Tseng W-YI (2010): IEEE transactions on medical imaging 29:1626–1635.

Yu T, Wang X, Li Y, Zhang G, Worrell G, Chauvel P, Ni D, Qiao L, Liu C, Li L, others (2018): High-frequency stimulation of anterior nucleus of thalamus desynchronizes epileptic network in humans. Brain 141:2631–2643.

